# Evolution and neutralization escape of the SARS-CoV-2 BA.2.86 subvariant

**DOI:** 10.1101/2023.09.08.23295250

**Authors:** Khadija Khan, Gila Lustig, Kajal Reedoy, Zesuliwe Jule, Cornelius Römer, Farina Karim, Yashica Ganga, Mallory Bernstein, Zainab Baig, Boitshoko Mahlangu, Anele Mnguni, Ayanda Nzimande, Nadine Stock, Dikeledi Kekana, Buhle Ntozini, Cindy van Deventer, Terry Marshall, Nithendra Manickchund, Bernadett I. Gosnell, Richard J. Lessells, Quarraisha Abdool Karim, Salim S. Abdool Karim, Mahomed-Yunus S. Moosa, Tulio de Oliveira, Anne von Gottberg, Nicole Wolter, Richard A Neher, Alex Sigal

## Abstract

Omicron BA.2.86 subvariant differs from Omicron BA.2 as well as recently circulating variants by over 30 mutations in the spike protein alone. Here we report on the first isolation of the live BA.2.86 subvariant from a diagnostic swab collected in South Africa which we tested for escape from neutralizing antibodies and viral replication properties in cell culture. BA.2.86 did not have significantly more escape than Omicron XBB.1.5 from neutralizing immunity elicited by infection of Omicron subvariants ranging from BA.1 to XBB, either by infection alone or as breakthrough infection in vaccinated individuals. Neutralization escape was present relative to earlier strains: BA.2.86 showed extensive escape both relative to ancestral virus in sera from pre-Omicron vaccinated individuals and relative to Omicron BA.1 in sera from Omicron BA.1 infected individuals. We did not observe substantial differences in viral properties in cell culture relative to XBB.1.5. Both BA.2.86 and XBB.1.5 produced infection foci of similar size, had similar cytopathic effect (both lower than ancestral SARS-CoV-2), and had similar replication dynamics. We also investigated the relationship of BA.2.86 to BA.2 sequences and found that the closest were BA.2 samples from Southern Africa circulating in early 2022. These observations suggest that BA.2.86 is more closely related to sequences from Southern Africa than other regions and so may have evolved there, and that evolution led to escape from neutralizing antibodies similar in scale to recently circulating strains of SARS-CoV-2.

---

The Omicron subvariant BA.2.86 is derived from the BA.2 subvariant but has over 30 mutations in spike relative to both BA.2 and recently circulating subvariants such as XBB.1.5 (Fig 1A), making its emergence a major concern since many of the mutations are predicted to confer escape from neutralizing antibodies (1). Levels of neutralizing antibodies have been found to strongly correlate with protection from symptomatic infection with SARS-CoV-2 (2). SARS-CoV-2 variant mutations occurring in the receptor binding domain and N terminal domain of spike tend to reduce the ability of antibodies elicited by previous infection or vaccination to neutralize SARS-CoV-2 (3-6), although protection from more severe disease may be partly maintained by conserved T cell responses (7).

**Figure 1:**
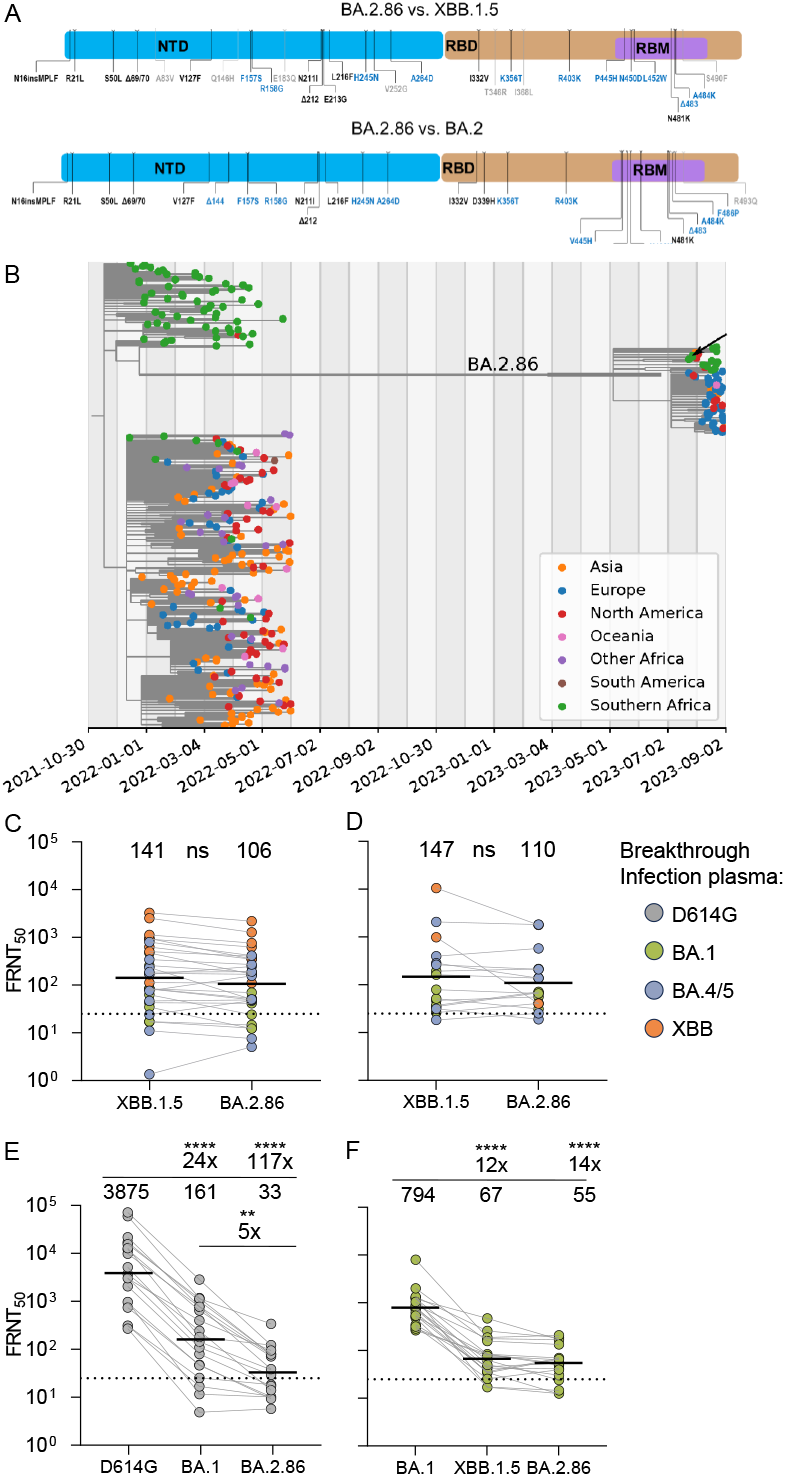
BA.2.86 evolution and neutralization escape. (A) Changes relative to Omicron XBB.1.5 and BA.2. (B) Phylogenetic analysis. BA.2.86 sequences form a distinct cluster separated from BA.2 sequences circulating late 2021/early 2022 by a long branch labeled BA.2.86. Outgrown sample marked by arrow. The BA.2.86 branch connects to samples with the mutations C26681T and C24378T (Spike: S939F) but lacks C9866T present in most BA.2 sequences. (C) Neutralization of BA.2.86 live virus versus XBB.1.5 by sera from vaccinated individuals with breakthrough BA.1/BA.4/BA.5/XBB family subvariant infection. Numbers above columns are group geometric mean titer FRNT_50_ and fold-change. (D) BA.2.86 versus XBB.1.5 neutralization by sera from unvaccinated individuals infected with Omicron BA.1/BA.4/BA.5/XBB family subvariants. (E) Neutralization of BA.2.86 versus ancestral D614G and BA.1 viruses by sera from vaccinated individuals collected before Omicron emergence. (F) Neutralization of BA.2.86 versus Omicron BA.1 by sera from vaccinated and unvaccinated BA.1 infected individuals. Significant p-values were ****p<0.0001, **p=0.0042 by the Wilcoxon Rank Sum test.

The Omicron BA.2.86 subvariant started to be identified by global genomic surveillance samples collected from 24 July 2023 onwards, but because of the reduced rate of surveillance the exact time when it started to spread is unclear. Likewise, it is unclear when and where it arose. BA.2.86 shares the synonymous mutation C26681T and the spike substitution S939F with BA.2 genomes sampled in South Africa in early 2022, while it lacks the mutation C9866T that is present in the great majority of BA.2 sequences sampled outside of Southern Africa (Fig 1B). Southern African sequences are also closely related to the putative ancestral sequence of BA.2.86. Of the 10 branches that emanate from the basal polytomy within BA.2.86, 8 are dominated by samples from Southern Africa (Fig 1B). Most sequences from the Northern Hemisphere fall into a second large polytomy designated as BA.2.86.1, separated from the basal polytomy by two mutations. Available BA.2.86 sequences differ from their most recent common ancestor (MRCA) by 1-7 mutations. Most samples were collected mid-August and have 3-5 mutations relative to the MRCA. SARS-CoV-2 accumulates about 15 mutations per year along acute transmission chains and we thus estimate that this subvariant started to spread about May 2023 (8). The estimate is corroborated by molecular clock analysis with TreeTime, which suggests an emergence date in early May with an uncertainty of about two months (Fig 1B).

The virus tested here is from a nasopharyngeal swab sample collected in Mpumalanga Province, South Africa on July 28, 2023 (Fig 1B, arrow). Sequence results were released on August 22, 2023 (GISAID accession GISAID accession EPI_ISL_18125249). Outgrowth to expand this virus was started on August 24, 2023, in the Vero-TMPRSS cell line, where two passages were performed (Materials and Methods). The sequence of the outgrown virus was deposited on GISAID (EPI_ISL_18226980) with no clear in-vitro sequence changes detected relative to the accepted Omicron BA.2.86 sequence.

Because the pressing concern is that BA.2.86 can escape current population immunity, we compared neutralization of BA.2.86 to XBB.1.5 using sera from individuals who were vaccinated and had breakthrough infection with an Omicron subvariant (Fig 1C, see Table S1 for participant details), or infected with an Omicron subvariant only (Fig 1D, see Table S2 for participant details). We found that there was no significant escape relative to XBB.1.5 in either of these groups. The exceptions were two unvaccinated participants who were infected during the XBB dominant period who did show substantial BA.2.86 escape relative to XBB.1.5 (Fig 1D and Table S2). However, it is difficult to determine if this is part of a pattern since we could not enroll more unvaccinated XBB subvariant or later subvariant infected participants. Some of the participants were people living with HIV (PLWH). In all except for one case HIV was suppressed with effective antiretroviral therapy. HIV status did not noticeably change the similar neutralization of XBB.1.5 and BA.2.86 (Fig S1).

Next, we examined if this variant evolved escape to neutralizing immunity relative to earlier SARS-CoV-2 strains. We checked neutralization of vaccinated individual sera collected pre-Omicron which we previously used to determine escape of the original Omicron BA.1 subvariant (3). Here, we found over 100-fold escape of BA.2.86 relative to ancestral SARS-CoV-2, 5-fold greater than observed for BA.1 (Fig 1E). We also tested for escape relative to Omicron BA.1 in people infected with BA.1. Here we again found extensive escape, 14-fold relative to BA.1. However, XBB.1.5 showed a similar, 12-fold escape (Fig 1F).

We then investigated whether there were any differences in focus formation - the number of cells infected by a single cell which formed an infected cell cluster. This is a measure of cell-to-cell spread of the virus (9). We also measured cytopathic effect and viral replication. We found that both BA.2.86 and XBB.1.5 made infection foci which were 5-fold and 4.5-fold smaller in area relative to those made by ancestral SARS-CoV-2 at 20 hours post-infection (Fig 2A, quantitation in Fig 2B). The same viral inoculum led to considerable cytopathic (CPE) effect by 72 hours in ancestral SARS-CoV-2 infected cells. Similar number of infection foci led to substantially less CPE at the same timepoint in both BA.2.86 and XBB.1.5 infections (Fig 2A). We also could not detect substantial differences in replication in cell culture between BA.2.86 and XBB.1.5 (Fig 2C).

**Figure 2:**
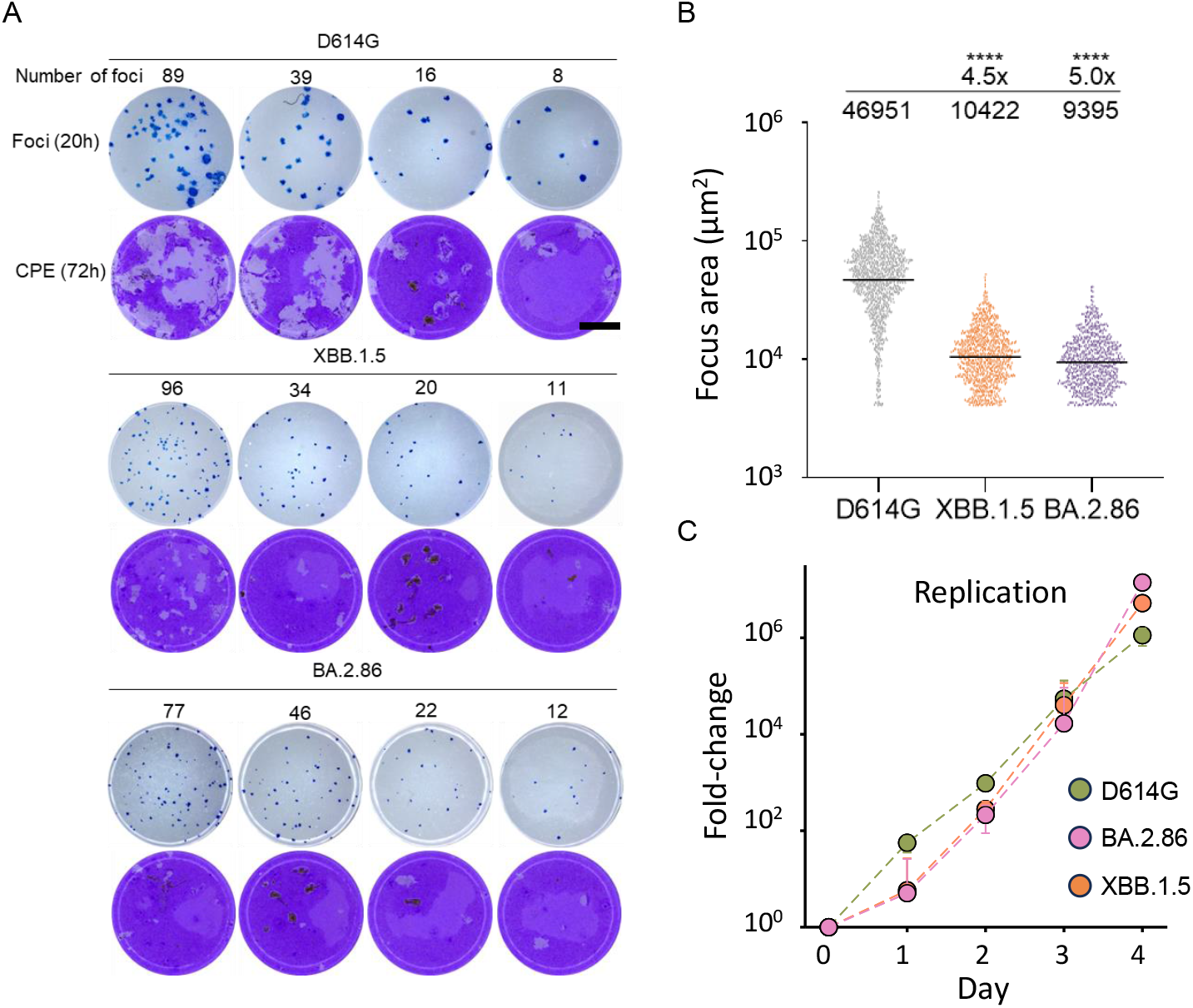
Omicron BA.2.86 replication and spread in cell culture. (A) Foci formed by ancestral D614G, XBB.1.5, and BA.2.86 at 20 hours post-infection on Vero-TMPRSS2 cells (rows 1, 3, 5) and cytopathic effect formed by the same viral inoculum on Vero-TMPRSS2 at 72 hours post-infection (rows 2, 4, 6). Rows 1 and 2 were infected with ancestral SARS-CoV-2, 3 and 4 with XBB.1.5, and 5 and 6 with BA.2.86. Foci number per well is indicated above each well. Bar is 2mm. (B) Quantitation of focus area for D614G, XBB.1.5, and BA.2.86. Significant p-values were ****p<0.0001 by the Wilcoxon Rank Sum test. (C) Fold-change in SARS-CoV-2 viral copies as determined using qPCR cycle threshold over 4 days of infection in H1299-ACE2 cells.

These results indicate that, although the Omicron BA.2.86 subvariant has evolved extensive escape from neutralizing antibodies, it is recognized by convalescent plasma to a similar degree as the XBB.1.5 subvariant. This similarity in recognition might explain the comparatively slow spread of the variant relative to the spread of BA.1 or BA.2 in 2021. These observations are similar to data reported over the past week showing limited or no escape of BA.2.86 relative to current circulating variants (10-12). Unlike the first study to report (11), we did not obtain data consistent with lower cellular infectivity of BA.2.86 relative to XBB.1.5, although this may be because of the different cellular assays used. A limitation of our data is that it is based on a single BA.2.86 isolate.

Our analysis suggests that BA.2.86 descends from viruses that circulated in early 2022 without any observed intermediates and only started to spread recently. There may be several explanations for the long period of evolution in the absence of population spread, including evolution in long-term SARS-CoV-2 infection during immunosuppression due to factors such as advanced HIV disease (13-15) as well as infection in an animal reservoir (15, 16).

While BA.2.86 may lead to new infections in the population, our data does not indicate that it is very different than SARS-CoV-2 strains already in circulation.

## Methods

### Informed consent and ethical statement

Blood samples and nasopharyngeal swab for ancestral D614G SARS-CoV-2 isolation were obtained after written informed consent from adults with PCR-confirmed SARS-CoV-2 infection who were enrolled in a prospective cohort study at the Africa Health Research Institute approved by the Biomedical Research Ethics Committee at the University of KwaZulu–Natal (reference BREC/00001275/2020). The Omicron/BA.1 and BA.2.86 was isolated from a residual swab sample with SARS-CoV-2 isolation from the sample approved by the University of the Witwatersrand Human Research Ethics Committee (HREC) (ref. M210752). The sample to isolate XBB.1.5 was collected after written informed consent as part of the COVID-19 transmission and natural history in KwaZulu-Natal, South Africa: Epidemiological Investigation to Guide Prevention and Clinical Care in the Centre for the AIDS Programme of Research in South Africa (CAPRISA) study and approved by the Biomedical Research Ethics Committee at the University of KwaZulu–Natal (reference BREC/00001195/2020, BREC/00003106/2021).

### Reagent availability statement

Viral isolates are available upon reasonable request. Sequences of isolated SARS-CoV-2 used in this study have been deposited in GISAID with accession numbers as follows:

| **Virus** | **GISAID** |
| --- | --- |
| **D614G** | EPI_ISL_602626.1 |
| **XBB.1.5** | EPI_ISL_17506815 |
| **BA.2.86** | EPI_ISL_18226980 |
| **BA.1** | EPI_ISL_7886688 |

Sequences used in phylogenetic analysis: GISAID Identifier: EPI_SET_230907xn, doi: 10.55876/gis8.230907xn. All genome sequences and associated metadata in this dataset are published in GISAID’s EpiCoV database. To view the contributors of each individual sequence with details such as accession number, Virus name, Collection date, Originating Lab and Submitting Lab and the list of Authors, see 10.55876/gis8.230907xn

Data Snapshot: EPI_SET_230907xn is composed of 347 individual genome sequences. The collection dates range from 2021-12-14 to 2023-08-30; Data were collected in 42 countries and territories; All sequences in this dataset are compared relative to hCoV-19/Wuhan/WIV04/2019 (WIV04), the official reference sequence employed by GISAID (EPI_ISL_402124). For more information see at https://gisaid.org/WIV04.

### Whole-genome sequencing and genome assembly

For the BA.2.86 swab sample, RNA was extracted on an automated Chemagic 360 instrument, using the CMG-1049 kit (Perkin Elmer, Hamburg, Germany). Libraries for whole genome sequencing were prepared using the Illumina COVIDseq Assay (Illumina Inc, San Diego, CA) and version 4 SARS-CoV-2 primer pools. Pooled PCR products were fragmented and tagged to adapter sequences. The adapter-tagged amplicons were purified and indexed using sets 1-4 of PCR indexes (Illumina). Libraries were quantified using a Qubit 4.0 fluorometer (ThermoFisher Scientific, Oregon, USA) using the Qubit dsDNA High Sensitivity assay according to the manufacturer’s instructions. Fragment sizes were analyzed using the TapeStation 4200 system (Agilent Technologies, Santa Clara, CA). Libraries were pooled and normalized to 4nM sample library with 2% PhiX spike-in. Libraries were loaded onto a 300-cycle NextSeq P2 Reagent Kit v2 and run on the Illumina NextSeq 1000/2000 instrument (Illumina). Sequencing data was analyzed using Exatype v4.1.5 (Hyrax Biosciences, Cape Town, South Africa) with default parameters (10% minimum prevalence to report variants, 80% minimum prevalence to include a variant in consensus sequence). Nextclade (v2.14.1) and Pangolin (v4.3, Pangolin-data v1.21) were used for clade and lineage assignments. Additionally Nextclade was used for visualization of the sequences and the identification of frameshifts. Unknown frameshifts were manually corrected using Aliview (v1.24). Outbreak.info was used to determine the prevalence of mutations.

For the BA.2.86 outgrowth sample, Oxford Nanopore sequencing was performed. RNA was manually extracted from either 200uL input volume using either the MagMAX(tm) Viral/Pathogen II Nucleic Acid Isolation Kit (Thermo Scientific) or from 140uL using the QIAamp Viral RNA Kit (Qiagen) as per the manufacturer’s protocols. All RNA extractions were measured using Qubit fluorimeter kits (Thermo Scientific). The cDNA synthesis was performed using LunaScript RT mastermix (New England BioLabs) followed by whole-genome multiplex PCR using the Midnight Primer pools v3 (EXP-MRT001, Oxford Nanopore) that produce 1,200-base-pair amplicons. The amplified products for each pool were combined and used to library preparation procedures using Oxford Nanopore Rapid Barcoding kit (SQK-RBK110.96, Oxford Nanopore). The barcoded samples were pooled and cleaned-up using magnetic beads and loaded on an R9.4.1 flow cell for 8-hours sequencing on a MinION device. The raw data was processed using Guppy basecaller and Guppy barcoder (Oxford Nanopore) for basecalling and demultiplexing. The final consensus sequences were obtained using the Genome Detective v2.64. The lineage assignment was determined using Nextclade.

### Phylogenetic analysis

We assembled a set of 280 BA.2 (Nextstrain clade 21L) sequences collected between November 2021 and June 2022 from data deposited on GISAID (17). BA.2.86 sequences were downloaded on September 7 2023 directly from GISAID. We excluded sequences with reversion mutations relative to BA.2, sequences flagged as poor quality by Nextclade (18), or sequences with less than 90% coverage of the reference. Sequences were pairwise aligned against Wuhan-Hu-1 using Nextclade. Terminals and gaps were masked as well as all suspected artefactual reversions to reference in BA.2.86 sequences. A tree was built using IQ-tree 2 (19) and postprocessed using a custom script to correct for incomplete merging of branches in large polytomies.

A time tree was inferred using TreeTime (20) using a clock rate of 0.0005 per site and year [Neher 2022]. The rate of the long branch between BA.2 and BA.2.86 was set to be 2 times the rate of the rest of the tree in line with previous observation that evolution is 2-fold accelerated along many long branches leading to distinct clades [Neher (2022)]. This acceleration is consistent with the dramatic enrichment of amino acid substitutions in the spike protein along the long branch leading to BA.2.86.

The phylogenetic workflow is available at github.com/neherlab/BA286. The repository contains a specific list of sites (config/mask.tsv) that are masked in individual sequences.

An interactive version of the phylogenetic tree is available at https://nextstrain.org/groups/neherlab/ncov/BA.2.86

### Cells

The VeroE6 cells expressing TMPRSS2 and ACE2 (VeroE6-TMPRSS2), originally BEI Resources, NR-54970 were used for virus expansion and all live virus assays excluding replication. The Vero-TMPRSS2 cell line was propagated in growth medium consisting of Dulbecco’s Modified Eagle Medium (DMEM) with 10% fetal bovine serum (Hyclone) containing 10mM of hydroxyethylpiperazine ethanesulfonic acid (HEPES), 1mM sodium pyruvate, 2mM L-glutamine and 0.1mM nonessential amino acids (Sigma-Aldrich). The H1299-E3 (H1299-ACE2, clone E3) cell line used in the replication assay was derived from H1299 (CRL-5803) and propagated in growth medium consisting of complete Roswell Park Memorial Institute (RPMI) 1640 with 10% fetal bovine serum containing 10mM of HEPES, 1mM sodium pyruvate, 2mM L-glutamine and 0.1mM nonessential amino acids.

### Virus expansion

All work with live virus was performed in Biosafety Level 3 containment using protocols for SARS-CoV-2 approved by the Africa Health Research Institute Biosafety Committee. VeroE6-TMPRSS2 cells were seeded at 4.5 × 10^5^ cells in a 6 well plate well and incubated for 18–20 hours pre-infection. After one Dulbecco’s phosphate-buffered saline (DPBS) wash, the sub-confluent cell monolayer was inoculated with 500μL with universal transport medium which contained the swab, diluted 1:2 with growth medium filtered through a 0.45μm and 0.22μm filters. Cells were incubated for 2 hours. Wells were then filled with 3 mL complete growth medium. After 3 days of infection (completion of passage 1 (P1)), supernatant was collected, cells were trypsinized, centrifuged at 300 rcf for 3 min and resuspended in 3mL growth medium. All infected cells and supernatant were added to VeroE6-TMPRSS2 cells that had been seeded at 1.5 × 10^5^ cells per mL, 20mL total, 18–20 hour earlier in a T75 flask for cell-to-cell infection. The coculture was incubated for 1 h and the flask was filled with 20mL of complete growth medium and incubated for 3 days. The viral supernatant from this culture (passage 2 (P2) stock) was used for experiments.

### Live virus focus forming assay and neutralization assay

VeroE6-TMPRSS2 cells were plated in a 96-well plate (Corning) at 30,000 cells per well 1 day pre-infection. Plasma was separated from EDTA-anticoagulated blood by centrifugation at 500 rcf for 10 min and stored at − 80 °C. Aliquots of plasma samples were heat-inactivated at 56 °C for 30 min and clarified by centrifugation at 10,000 rcf for 5 min. Virus stocks were used at approximately 50-100 focus-forming units per microwell and added to diluted plasma in neutralization assays. Antibody–virus mixtures were incubated for 1 h at 37 °C, 5% CO_2_. Cells were infected with 100 μL of the virus–antibody mixtures for 1 h, then 100 μL of a 1X RPMI 1640 (Sigma-Aldrich, R6504), 1.5% carboxymethylcellulose (Sigma-Aldrich, C4888) overlay was added without removing the inoculum. Cells were fixed 20 h post-infection using 4% PFA (Sigma-Aldrich) for 20 min. Foci were stained with a rabbit anti-spike monoclonal antibody (BS-R2B12, GenScript A02058) at 0.5 μg/mL in a permeabilization buffer containing 0.1% saponin (Sigma-Aldrich), 0.1% BSA (Sigma-Aldrich) and 0.05% Tween-20 (Sigma-Aldrich) in PBS. Plates were incubated with primary antibody for 2 h at room temperature with shaking, then washed with wash buffer containing 0.05% Tween-20 in PBS. Secondary goat anti-rabbit HRP conjugated antibody (Abcam ab205718) was added at 1 μg/mL and incubated for 2 h at room temperature with shaking. TrueBlue peroxidase substrate (SeraCare 5510-0030) was then added at 50 μL per well and incubated for 20 min at room temperature. Plates were imaged in an ImmunoSpot Ultra-V S6-02-6140 Analyzer ELISPOT instrument with BioSpot Professional built-in image analysis (C.T.L) which was also used to quantify areas of individual foci.

### Plaque Assay

VeroE6-TMPRSS2 cells were plated in a 96-well plate (Corning) at 30,000 cells per well 1 day pre-infection. Virus stocks were used at the focus-forming units per microwell shown in Figure 2 and added cells, incubated for 1 h at 37 °C, 5% CO2. Following incubation, 100μL of a 1X RPMI 1640 (Sigma-Aldrich, R6504), 1.5% carboxymethylcellulose (Sigma-Aldrich, C4888) overlay was added without removing the inoculum. Cells were fixed 72 hours post-infection using 4% PFA (Sigma-Aldrich) for 20 min. The fixed cells were washed with distilled water and stained with 30 μL/well of a 0.5% crystal violet solution (Sigma-Aldrich, 61135).

### Cycle threshold values for SARS-CoV-2 RNA copies

SARS-CoV-2 cycle threshold (Ct) quantification was performed from sample supernatant at an accredited diagnostic laboratory (Molecular Diagnostic Services, Durban, South Africa). Samples were extracted using a guanidine isothiocyanate/ magnetic bead based method with the NucliSense (Biomerieux) extractor of the KingFisher Flex 96 (Thermo Fisher). RT-qPCR was performed using the Seegene Allplex 2019 nCoV assay with the Bio-Rad CFX96 real-time PCR instrument as per the kit instructions. RNase P is used as the internal housekeeping gene to monitor extraction and assay efficiency. The kit targets the E, N and R genes of SAR CoV-2. Run calls and interpretation is performed by the Seegene Viewer software. Fold-change was calculated as FC = 2^((mean(Ct input) – Ct sample)^

### Statistics and fitting

All statistics and fitting were performed using custom code in MATLAB v.2019b. Neutralization data were fit to:

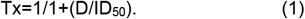

Here Tx is the number of foci at plasma dilution D normalized to the number of foci in the absence of plasma on the same plate. ID_50_ is the plasma dilution giving 50% neutralization. FRNT_50_ = 1/ID_50_. Values of FRNT_50_ <1 are set to 1 (undiluted), the lowest measurable value. We note that the most concentrated plasma dilution was 1:25 and therefore FRNT_50_ < 25 were extrapolated.

## Data Availability

All data produced in the present study are available upon reasonable request to the authors

## Acknowledgements

This study was supported by the Bill and Melinda Gates award INV-018944, Wellcome Trust Award 226137/Z/22/Z, University of Washington Arboviral Research Network (UWARN) Subaward #UWSC14272 and the South African Medical Research Council to AS. We thank the originating and submitting labs of the SARS-CoV-2 sequences used in this study for timely sharing of their data (EPI_SET_230907xn). We particularly thank Amos Adler and his team and the Danish COVID-19 Genomics Consortium, whose surveillance efforts have led to the detection of BA.2.86. We thank github user Silcn and Andrew Rambaut for discussions of phylogenetic aspects and sequence quality.

## Figure Legends

**Figure S1:**
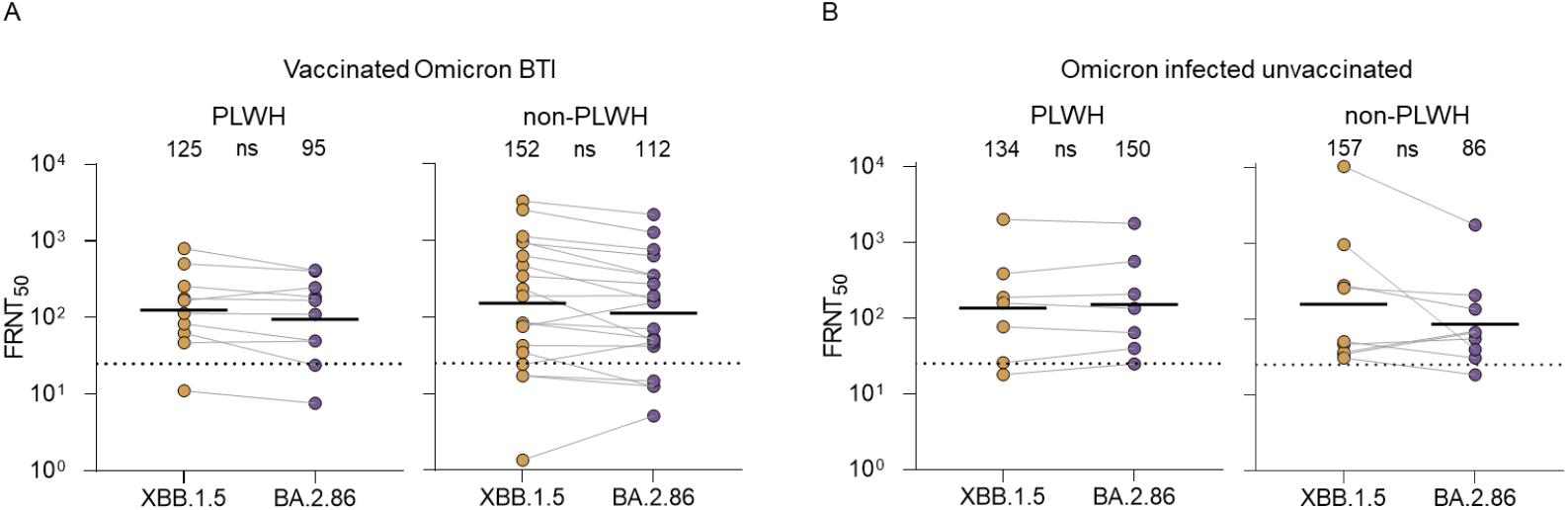
Neutralization escape of Omicron BA.2.86 relative to XBB.1.5 by HIV status. (A) Neutralization of XBB.1.5 and BA.2.86 in vaccinated individuals with breakthrough Omicron variant infection. Y-axis is neutralization as FRNT_50_ and numbers above each column are geometric mean titer (GMT) FRNT_50_ for the group. Sera from people living with HIV (PLWH) are on the left panel and HIV negative participants on the right. (B) Neutralization of XBB.1.5 and BA.2.86 in unvaccinated individuals infected with Omicron subvariants.

**Table S1:**
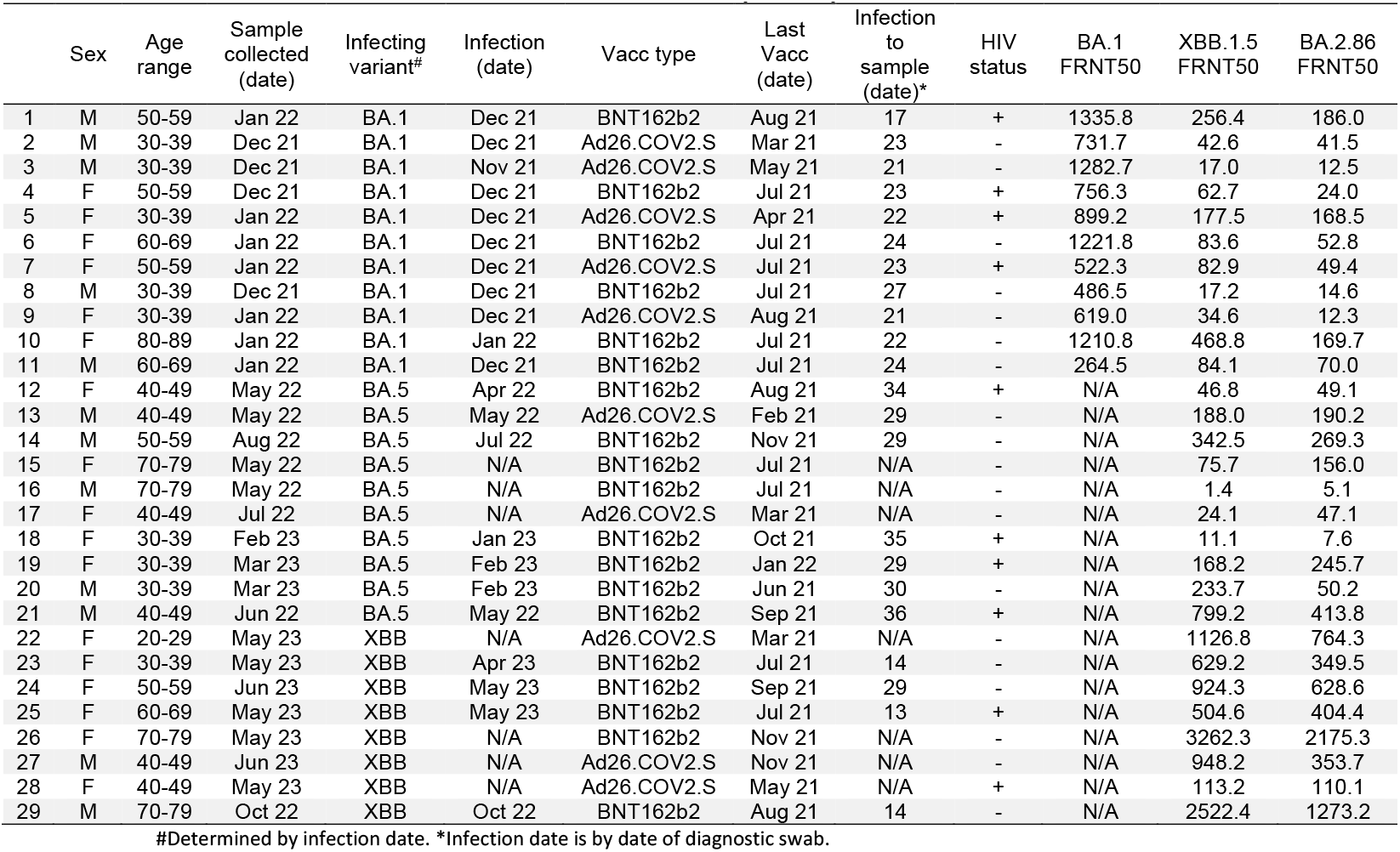
Vaccinated with Omicron BTI participants.

**Table S2:** Unvaccinated with Omicron infection participants.

|  | Sex | Age range | Sample collected (date) | Infecting Variant<sup>#</sup> | Infection Date<sup>*</sup> | Infection to sample (date) | HIV status | BA.1 FRNT50 | XBB.1.5 FRNT50 | BA.2.86 FRNT50 |
| --- | --- | --- | --- | --- | --- | --- | --- | --- | --- | --- |
| **1** | F | 30-39 | Jan 22 | BA.1 | Dec 21 | 23 | + | 7963.06 | 187.34 | 207.40 |
| **2** | F | 20-29 | Dec 21 | BA.1 | Dec 21 | 21 | - | 540.03 | 36.06 | 68.98 |
| **3** | F | 20-29 | Dec 21 | BA.1 | Dec 21 | 21 | - | 345.02 | 45.90 | 54.60 |
| **4** | F | 50-59 | Jan 22 | BA.1 | Dec 21 | 31 | + | 290.00 | 76.89 | 64.08 |
| **5** | F | 60-69 | Jan 22 | BA.1 | Dec 21 | 17 | - | 826.48 | 34.08 | 66.15 |
| **6** | F | 50-59 | Jan 22 | BA.1 | Dec 21 | 27 | + | 316.99 | 25.57 | 39.76 |
| **7** | F | 50-59 | Jan 22 | BA.1 | Dec 21 | 21 | - | 1046.76 | 49.91 | 30.38 |
| **8** | F | 50-59 | Jan 22 | BA.1 | Dec 21 | 31 | + | 2001.21 | 157.74 | 133.82 |
| **9** | F | 30-39 | May 22 | BA.5 | Mar 22 | 78 | + | N/A | 17.9 | 24.6 |
| **10** | M | 40-49 | Jun 22 | BA.5 | May 22 | 35 | + | N/A | 384.2 | 558.9 |
| **11** | F | 50-59 | Aug 22 | BA.5 | Jul 22 | 30 | - | N/A | 30.5 | 18.3 |
| **12** | M | 60-69 | Aug 22 | BA.5 | Jul 22 | 27 | - | N/A | 254.3 | 203.7 |
| **13** | F | 40-49 | Feb 23 | BA.5 | Jan 23 | 33 | - | N/A | 275.5 | 134.0 |
| **14<sup>\*\*</sup>** | F | 30-39 | Mar 23 | BA.5 | Mar 23 | 17 | + | N/A | 2003.2 | 1769.6 |
| **15** | M | 50-59 | May 23 | XBB | N/A | N/A | - | N/A | 10239.0 | 1735.9 |
| **16** | M | 60-69 | Apr 23 | XBB | Apr 23 | 28 | - | N/A | 955.9 | 39.2 |

#Determined by infection date. \*Infection date is by date of diagnostic swab. \*\*Viremic. HIV viral load = 2383 copies/mL.

**Table S3:** BNT162b2 Vaccinated pre-Omicron participants.

|  | Sex | Age range | Sample collected (date) | Last Vacc. (date) | Vacc. to sample (days) | HIV status | D614G FRNT50 | BA.1 FRNT50 | BA.2.86 FRNT50 |
| --- | --- | --- | --- | --- | --- | --- | --- | --- | --- |
| **1** | F | 40-49 | Sep 21 | Aug 21 | 32 | + | 942.3 | 10.9 | 9.6 |
| **2** | F | 40-49 | Oct 21 | Sep 21 | 34 | + | 16919.6 | 984.1 | 76.0 |
| **3** | F | 60-69 | Oct 21 | Aug 21 | 63 | - | 11788.5 | 621.0 | 68.1 |
| **4** | M | 40-49 | Oct 21 | Oct 21 | 18 | + | 9629.4 | 749.5 | 36.7 |
| **5** | M | 70-79 | Aug 21 | Jun 21 | 38 | - | 5025.0 | 137.8 | 35.6 |
| **6** | F | 60-69 | Jul 21 | Jul 21 | 11 | - | 301.1 | 16.0 | 9.9 |
| **7** | M | 70-79 | Jul 21 | Jul 21 | 11 | - | 720.2 | 45.3 | 8.8 |
| **8** | M | 30-39 | Aug 21 | Jul 21 | 15 | + | 893.5 | 44.2 | 9.7 |
| **9** | F | 70-79 | Jul 21 | Jul 21 | 10 | - | 300.5 | 24.2 | 17.3 |
| **10** | F | 30-39 | Jul 21 | Jul 21 | 11 | + | 2002.2 | 75.8 | 16.2 |
| **11** | F | 20-29 | Nov 21 | Oct 21 | 31 | - | 3625.0 | 234.5 | 13.3 |
| **12** | M | 20-29 | Nov 21 | Oct 21 | 30 | - | 2963.4 | 104.0 | 27.7 |
| **13** | F | 60-69 | Aug 21 | Jul 21 | 28 | - | 59258.6 | 1143.2 | 74.3 |
| **14** | M | 60-69 | Aug 21 | Jul 21 | 26 | - | 71412.3 | 2779.6 | 117.2 |
| **15** | F | 40-49 | Nov 21 | Oct 21 | 33 | + | 15276.3 | 1104.9 | 327.6 |
| **16** | M | 50-59 | Oct 21 | Sep 21 | 30 | - | 21376.2 | 759.2 | 72.7 |
| **17** | F | 50-59 | Nov 21 | Oct 21 | 13 | - | 2915.2 | 385.3 | 90.7 |
| **18** | F | 60-69 | Nov 21 | Nov 21 | 14 | + | 12699.9 | 174.2 | 117.7 |
| **19** | F | 30-39 | Aug 21 | Jul 21 | 34 | + | 258.6 | 4.6 | 5.4 |

